# Effect of results-based financing on selected maternal and child health indicators in Mayuge District, Uganda: a comparative interrupted time series analysis

**DOI:** 10.64898/2026.11.04.26367302

**Authors:** Haji Eboga, Yovani Lubaale, Faith Oguttu, Ritah Nantale, Prince Tsekpetse, Brian Tonny Makoko, Elizabeth Kawala, Richard Mugahi, Andrew Kazibwe, David Okia, Milton W Musaba, Dan Kibuule, David Mukunya

## Abstract

**Background:** Uganda continues to experience a high burden of maternal and child morbidity and mortality, partly due to suboptimal utilization and quality of reproductive, maternal, newborn, and child health services. To address these challenges, the Ministry of Health of Uganda, with World Bank funding, implemented results-based financing (RBF) through the Uganda Reproductive, Maternal, and Child Health Services Improvement Project (URMCHIP) in selected districts. Although RBF has been implemented for nearly a decade, evidence on its impact on maternal and child health service indicators in Uganda remains limited. This study assessed the effect of RBF on selected maternal and child health indicators in Mayuge District, Uganda.

**Methods:** We retrospectively reviewed the records between July 2018 and June 2019 as the pre-intervention period, July 2019 to June 2021 as the intervention period, and July 2021 to June 2023 as the post-intervention period. We conducted multiple-group interrupted time-series analyses to compare trends in selected maternal and child health indicators between four intervention and three non-intervention health facilities.

**Results:** After the implementation of RBF, the number of births in the intervention facilities increased by 1.67 births per month (95% CI [0.19, 3.16]) as compared to the control facilities. The number of women receiving modern family planning methods in intervention facilities increased by 34.1 women per month (CI [25.8, 42.4] as compared to the control facilities. The number of children under five years seen at the outpatient department slightly decreased by −0.24 children per month (CI [−9.58, 9.10]).

There was a decline in facility-based births and the number of clients receiving modern family planning methods following the discontinuation of results-based financing in the intervention health facilities.

**Conclusions:** There was an improvement in selected maternal and child health indicators in the intervention health facilities following the implementation of RBF, with a subsequent decline in facility-based births and the number of clients receiving modern family planning methods after RBF was discontinued. We recommend that policymakers consider allocating additional funds towards RBF to improve health service delivery.

## Background

Results-based financing (RBF) in healthcare refers to financial or material rewards given to a provider of health services after assessment of performance and verification of the attainment of a given target for selected health indicators [1]. Results-based financing links incentives to targeted health services, which in turn motivates health workers to improve the quality and quantity of the health services delivered [2]. Result-based financing is one of the strategies adopted by different countries to enhance the performance of health systems and achieve universal health coverage [3]. Emphasis is placed on selected health indicators, which the donors assume will have the most impact on the well-being of the most vulnerable populations, such as women, pregnant mothers, and children [4].

Low- and middle-income countries account for 95% of maternal deaths [5], 80% of the unmet need for modern contraception [6] and 90% of under-five mortality [7]. Uganda’s maternal mortality rate is 189 maternal deaths per 100,000 live births, under-five mortality is at 52 deaths per 1000 live births, and a 42% unmet need for modern contraceptive use [8]. One of the reasons for the persistently high burden of maternal, newborn, and child morbidity and mortality is the low utilization of reproductive, maternal, newborn and child health services [9, 10]. Services provided in public health facilities are perceived as low quality due to inadequate staff, staff absenteeism, limited working hours, poor attitude of health workers, and stockouts of drugs and other commodities [9]. Effective implementation of health financing models such as RBF has been shown to improve the performance of health workers and enhance the quality of health services provided [11, 12].

Uganda’s Ministry of Health, with funding from the World Bank, launched the Uganda Reproductive, Maternal, and Child Health Services Improvement Project (URMCHIP) in 2016 [4, 13]. The URMCHIP implemented a results-based financing mechanism to provide a flexible stream of health financing to lower health facilities.

Through RBF, the Ministry of Health, together with district local governments, developed scoring criteria, allocated funds and supervised health facility teams to implement local interventions aimed at improving maternal, newborn and child health outcomes [3, 14].

Few studies have reported the effect of RBF on service delivery and uptake in different regions of the country [14-16].

There have been differing reports on the impact of RBF on improving maternal, newborn, and child health services in low- and middle-income countries [3, 17, 18]. Further studies are needed to fully understand the impact of RBF on maternal, newborn, and child health services in our setting. Our study aimed to assess the effect of results-based financing on selected maternal, newborn, and child health indicators at four health facilities that implemented the intervention in comparison with three control health facilities in rural Eastern Uganda.

## Methods

### The intervention

The results-based financing project provided additional funding to selected health facilities to improve service delivery and increase the utilization of selected reproductive, maternal, newborn, and child health (RMNCH) services. Participating health facilities were granted autonomy to use additional RBF revenue, above their regular primary healthcare (PHC) non-wage allocations, to enhance performance. They could use their RBF funding for providing bonuses to health workers, procuring supplementary medicines and supplies, medical equipment, renovating their facilities, recruiting additional staff, facilitating Village Health Teams for community mobilization, and more [14].

Funds were intended to be released quarterly directly to the health facility accounts. The funds were withdrawn by the health facility in-charge, with 30% allocated to reward health workers. A rewards committee assessed individual health workers based on their performance against the RBF output indicators and allocated the staff incentives accordingly. The remaining 70% of the funds were used to improve service delivery at the health facility, including the purchase of drugs, hiring of additional staff, conducting outreach activities, purchasing equipment and instruments, renovations, and construction of new infrastructure.

The RBF output indicators were rewarded at the following rates: children under 5 years of age seen at the outpatient department (OPD), US$0.30 per child; health facility birth, US$8.10 per birth; and modern contraceptive use, US$5.40 per user.

### Study design

We used a quasi-experimental study design to assess the effect of RBF on the uptake of modern family planning methods, the number of children under five attending outpatient departments, and the number of women giving birth at health facilities. We retrospectively reviewed records from health facilities for patients seen during three periods: the pre-intervention period (July 2018–June 2019), the intervention period (July 2019–June 2021), and the post-intervention period (July 2021–June 2023).

Allocation to the intervention and control groups was determined by the Ministry of Health using predefined quality-of-care performance criteria [19]. Health facilities that achieved a quality-of-care assessment score of 70% or higher were selected to participate in the RBF scheme, while facilities that did not meet the required threshold served as controls. Four health center IIIs received RBF and three health center IIIs served as control facilities.

### Study setting

This study was conducted in Mayuge District. Mayuge district is located in Eastern Uganda, 120 km from Kampala. Mayuge has a total population of 562,000 [20]. At the time of RBF implementation, the district was served by one district hospital, three health center IVs, and seven health center IIIs. The district is characterised by poor maternal and child health outcomes. According to DHIS-2 Mayuge data, the maternal mortality ratio was 41 deaths per 100,000 live births, the infant mortality rate was 94 deaths per 1,000 live births, and the under-5 mortality rate was 123 deaths per 1,000 live births. The district-level rates for infant and under-5 mortality are considerably higher than the national rates of 36 and 52 deaths per 1,000 live births, respectively [21]. Institutional births are estimated at 41%, and unmet need for family planning at 47%, which is much lower than the national target. Inadequate staffing, constant absenteeism from duty by health workers, and poor staff motivation contribute to these poor outcomes. (DHIS-2 Mayuge). As an intervention to address the above health challenges, results-based financing was introduced and implemented by the Ministry of Health in four out of seven health center IIIs in Mayuge from 2019/2020 to 2020/2021 to improve performance.

### Study population

We reviewed records from four health centers that received result-based financing as the intervention group and three health centers that did not receive result-based financing in the financial years 2018/2019 to 2022/2023 as the control group. The HC IIIs in the intervention group included Malongo HC III, Buwaiswa HC III, Baitambogwe HC III, and Wabulungu HC III, and in the control group were Busaala HC III, Jaguzi HC III, and Masolya HC III.

### Sampling and Sample size

We reviewed all records of the selected monthly maternal and child health indicators from July 2018 to June 2023 from the seven health centers included in the study. All health center IIIs that existed at the beginning of the RBF project were selected.

Belonging to the control or intervention arm was predetermined at the time of RBF implementation by the Ministry of Health; we did not reallocate any health facility.

### Study variables

Results-based financing was the independent variable, while the number of children under five years old attending the outpatient department, the number of women using modern family planning, and the number of women giving birth at the health facility were the dependent variables.

### Data collection

Patient data were collected in registers by healthcare workers and then entered into the District Health Information System 2 by a data officer on a weekly basis. Data were downloaded from the District Health Information System 2 (DHIS 2) software, into a Microsoft excel form. For this study, the data were accessed on 15^th^ January 2024. Data were anonymized to remove individual participant identifiers, and a data abstraction tool was used to extract data on selected maternal, newborn, and child indicators. Data from the District Health Information System 2 (DHIS2) were cross-verified through physical register checks at the health facilities.

### Statistical analysis

Data were analyzed using Stata version 15 (StataCorp, College Station, TX, USA). We compared the data for three periods: the pre-intervention period (July 2018 to June 2019), the intervention period (July 2019 to June 2021) and the post-intervention period (July 2021 to June 2023).

We conducted multiple-group interrupted time series analyses (ITSA) to compare the trends of the different outcomes (number of women receiving modern family planning monthly, number of women giving birth at the health facility monthly, number of children under five years seen at the Outpatient Department (OPD) monthly) before, during, and after receiving result-based financing. We fitted different models for each outcome. We compared the trend in the intervention health facilities (four health facilities) to the trend in the control health facilities (three non-intervention facilities).

We then conducted a time series decomposition for the time before and after receiving result-based financing to identify trends, seasonality, and irregular components for each model.

This model assumes that the relationship between the dependent variables (family planning, OPD attendance, and institutional birth) and the exposure (result-based financing) is linear [22]. OLS regression was the most suitable because the result-based financing intervention and indicators were adequately captured by a linear model. We then interpreted the regression coefficient to understand the change in the level and trend after the intervention. Autocorrelation checks allowed us to decide on our OLS model (Newey with lag (1) specified).

Time series graphs were generated for each indicator, and we measured the change in the level (intercept) and slope (trend) of each indicator, before and after the intervention. We also used statistical techniques such as segmented regression analysis and interrupted time series analysis. We then determined the statistical significance of the intervention’s effect on the level and slope of the indicators.

## Results

### Comparison of the effect of result-based financing on the number of women using modern family planning methods between intervention and control health facilities

After the inception of results-based financing, the slope of the intervention group increased by 33.5 family planning users per month (p ≤ 0.001, 95% CI [27.6, 39.5]), while the change in slope in the control group was not statistically significant (p = 0.852, 95% CI [−6.32, 5.20]). The difference in slope change between the two groups was 34.1 users per month (p ≤ 0.001, 95% CI [25.8, 42.4]). The intervention group showed an increase of 34.1 new family planning uptakes monthly, compared with the control group.

After RBF was discontinued, the slope of the intervention group was −14.6 births per month (p=0.026, 95% CI [−27.4, −1.8]). In the control group, there was hardly any change in the number of family planning uptakes per month (0.2) after result-based financing stopped. p=0.956, 95% CI [−7.7, 8.2], indicating that the intervention group showed a decrease of 14.8 in family planning uptake per month, compared to the control group, when result-based financing stopped (p=0.054, 95% CI [−29.9, 0.28]). This is illustrated in figure 1.

**Figure 1.**
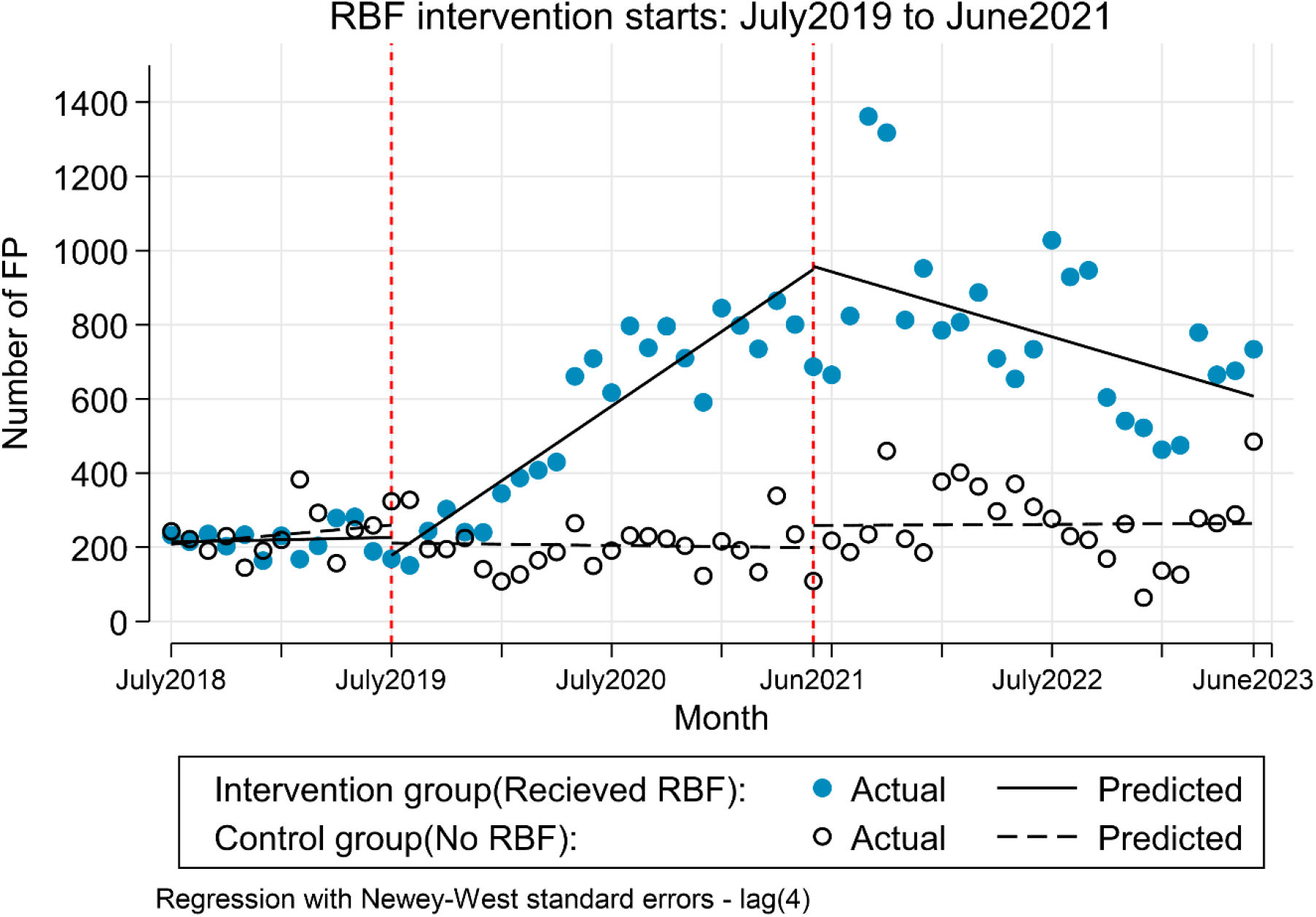
ITSA graph comparing number of women receiving modern family planning methods in health facilities that received RBF compared to those that did not receive RBF.

### Comparison of effect of result-based financing on number of women giving birth at the health facilities between intervention and control health facilities

After result based financing was started, the slope of the intervention group was 1.76 births per month (p=<0.008, 95% CI [0.48, 3.05]), and the change in trend in the control group showed no statistical significance (p=0.81, 95% CI [−0.66, 0.84]). The difference between the two groups was 1.67 births per month (p=< 0.028, 95% CI [0.19, 3.16]). The intervention group showed an increase of 1.67 new births monthly compared with the control group.

After RBF was stopped, the slope of the intervention group was −0.96 births per month (p=0.213, 95% CI [−2.47, 0.56]). In the control group, there was hardly any change in the number of births per month (0.35) after result-based financing discontinued. p=0.464, 95% CI [−1.29, 0.59]. The intervention group showed a decrease of 0.61 births per month compared to the control group when result-based financing stopped (p=0.501, 95% CI [−2.39, 1.18]). This is illustrated in Figure 2.

**Figure 2.**
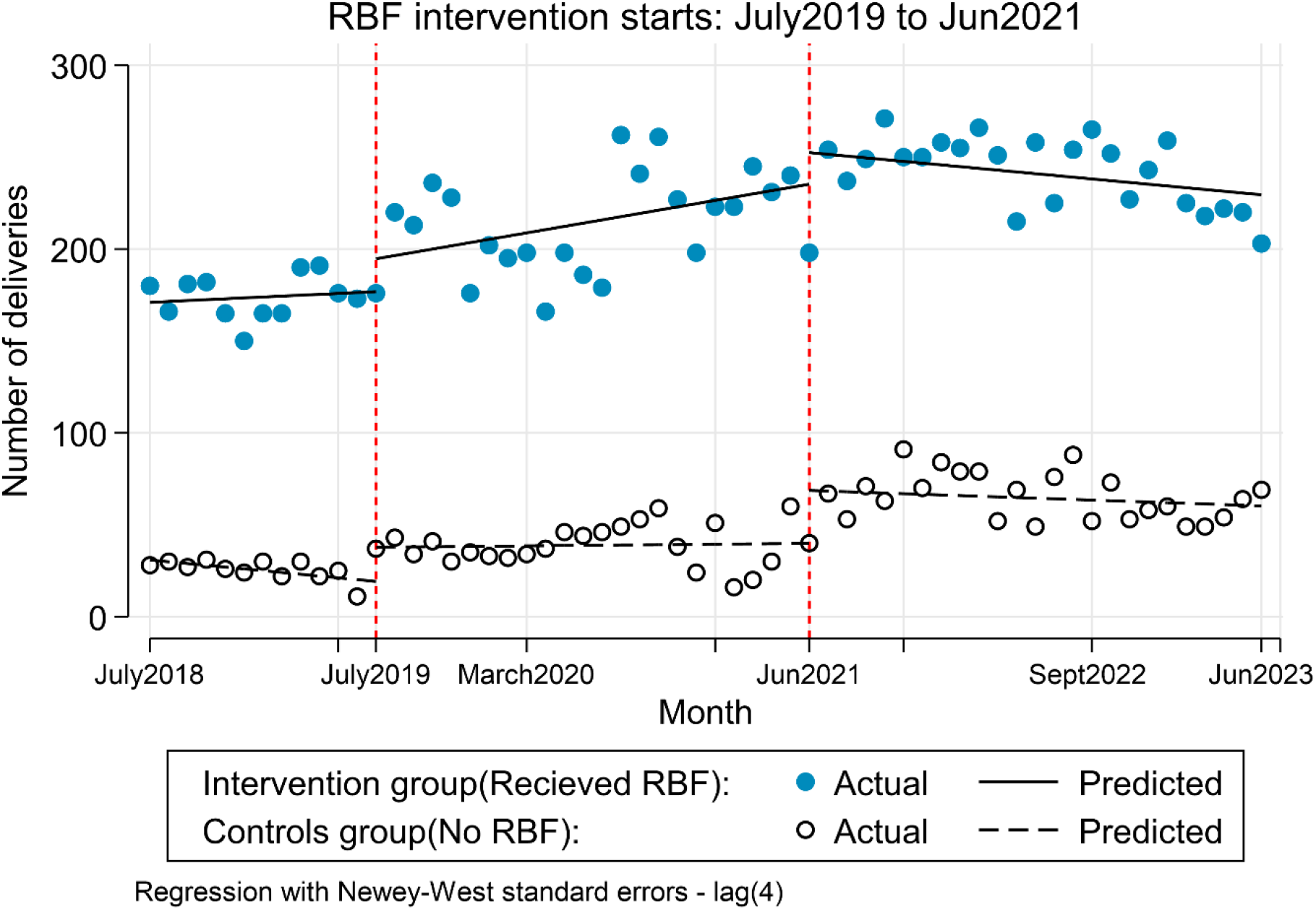
ITSA graph comparing number of women giving birth at health facilities that received RBF compared to those that did not receive RBF.

### Comparison of effect of result-based financing on number of children under five years attending the outpatient department between intervention and control health facilities

After result based financing was started, the slope of the intervention group was −0.24 children per month (p=0.960, 95% CI [−9.58, 9.10]), and the change in trend in the control group showed statistical significance (17.31, p<0.001; 95% CI [11.60, 23.02]). The difference between the two groups were −17.55 children per month (p=0.002, 95% CI [−28.49, −6.60]), indicating that the intervention group showed a decrease of 17.55 new OPD under five monthly visits, compared with the control group.

After RBF was stopped, the slope of the intervention group was 7.59 children per month (p=0.28, 95% CI [−6.24, 21.43]). In the control group, the slope of the number of OPD under five years per month was 6.68, after result-based financing stopped. p=0.07, 95% CI [−0.67, 14.02], indicating that the intervention group showed a decrease in 0.92 children per month compared to the control group when result-based financing stopped (p=0.91, 95% CI [−14.74, 16.58]). However, the change was not significant, this is shown in Figure 3.

**Figure 3.**
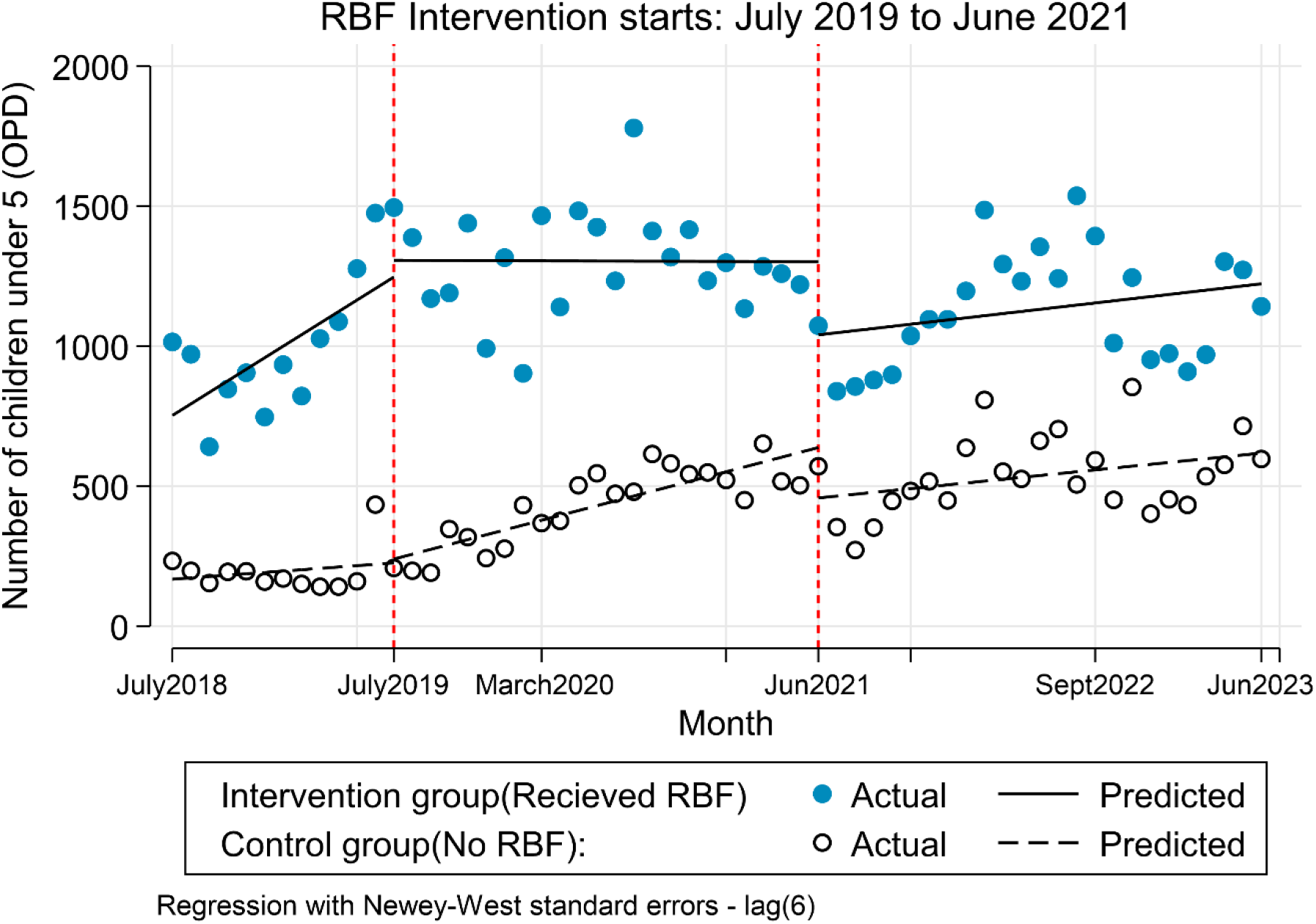
ITSA graph comparing number of children attending the outpatient department at health facilities that received RBF compared to those that did not receive RBF.

## Discussion

Our study found that RBF significantly improved the uptake of modern family planning and the number of women giving birth at the health facilities but did not significantly change the number of children under five years attending the outpatient department in the intervention health facilities.

### Effect of RBF on uptake of modern family planning methods

The number of women taking up modern family planning increased by nearly five times after the implementation of the RBF. This number gradually decreased after RBF was discontinued. There was no change in the number of women using modern family planning methods in control health facilities. Our findings are similar to those of studies conducted in Cameroon and Congo, which also found an increased uptake of family planning among women in communities and health facilities where RBF was implemented [23-25].

The increase in the uptake of family planning may be attributed to family planning outreaches and intensive community sensitization about family planning services, which was supported by the RBF fund [14]. Improved availability of family planning methods, coupled with motivated healthcare workers providing services, could have encouraged more women to utilize family planning methods.

The decline in the number of women taking up family planning supports our findings and suggests that withdrawal of incentives could have demotivated health workers. This is similar to findings from Egypt, which reported a decline in the uptake of family planning after the discontinuation of performance-based financing [26]. The lack of funds to conduct family planning outreaches, intensive community mobilization, and sensitization about family planning can explain why the number of women taking up modern family planning methods reduced after RBF was stopped. Our findings suggest that, given adequate funding, the challenge of unmet need for family planning can be effectively addressed. We encourage stakeholders to prioritize funding towards family planning.

### Effect of RBF on number of women giving birth at health facilities

During the intervention period, the intervention health facilities noted an increase of 1.67 new births monthly compared to the control health facilities. After the RBF was stopped, the intervention health facilities noted a decline in the number of births per month. The change in the number of new births in the control health facilities was not significant. Our findings are similar to studies done in Burkina Faso and Congo which also reported increased number of women giving birth at health facilities where RBF was implemented [25, 27].

This is attributed to intensive community mobilization about the value of giving birth in health facilities, availability of drugs and supplies, and support from RBF funds [14]. This attracted mothers to come and deliver from health facilities. This can also be explained by the possibility that healthcare workers were motivated to provide improved antenatal, intrapartum, and postpartum care, thus encouraging more women to give birth at health facilities.

There was a reduction in the number of women giving birth in the intervention health facilities compared with the control health facilities during the post-intervention period. This can be attributed to reduced community mobilization and sensitization about the value of giving birth in health facilities and stockouts of drugs and supplies that were previously supported by RBF funds.

### Effect of RBF on number of children under five years seen at the outpatient (OPD)

Compared to the pre-intervention period, facilities that received RBF registered an absolute increase in the number of children under five, attending the OPD clinic, compared to the control facilities. However, the rate of increase in the number of children who attended the OPD was not significant. These findings are similar to those of studies conducted in Burkina Faso, Congo, and Cameroon, which did not find any significant effects of RBF on the number of children seen at the health center[28] or care given to children under 5 years of age [23, 25, 29]. Under five, OPD attendance was among the least paid (0.3 USD), which could have caused health workers to concentrate on indicators that were paid for highly like institutional birth (8.3 USD). We recommend further studies to assess why RBF did not significantly affect the number of children under five years of OPD attendance

There was an increase in the number of children attending the control health facilities during the intervention. Unfortunately, our study did not explore any other programs that could have been running at the intervention health facilities to improve health service delivery.

### Strengths and limitations

Our study was able to assess selected maternal, newborn, and child health services over three periods: pre-implementation, during implementation, and after implementation of the RBF programme. The use of a controlled interrupted time-series design also allowed us to assess changes in the level and trend of the selected health indicators over time in the intervention and control facilities.

We did not collect data from the different health facilities regarding other programmes or interventions that may have been implemented concurrently to promote maternal, newborn, and child health services. Therefore, we cannot establish a direct causal link between RBF and the observed changes in the health indicators. We also collected retrospective data, which may not accurately reflect the current situation at the health facilities.

Another limitation is that the intervention and control facilities were not matched. Facilities that implemented RBF were selected by the Ministry of Health based on predefined criteria, including a minimum level of performance on the quality-of-care assessment. Only high-volume facilities were considered for the programme. Consequently, the intervention and control facilities may have differed in their baseline characteristics and performance, introducing the possibility of selection bias. Although the controlled interrupted time-series design allowed us to compare changes in outcome levels and trends between the intervention and control facilities over time, it cannot eliminate bias resulting from the non-random selection of facilities. The findings should therefore be interpreted with caution when attributing observed changes specifically to RBF.

The quality of the routine health facility data is another potential limitation. Because RBF payments were linked to reported performance, health workers may have inflated the number of patients attended to or services provided to increase the funds allocated to their facilities. Although we could not independently verify all reported figures, reporting bias cannot be ruled out, and the findings should therefore be interpreted with caution.

## Conclusion

Overall, an improvement was seen in maternal, newborn, and child services in the intervention health facilities during the intervention period, and a decline was noted following the termination of RBF. To ensure sustainability, we recommend that policymakers plan for the allocation of additional funds to RBF to promote better service delivery in health facilities.

## Data Availability

All data produced in the present study are available upon reasonable request to the authors

## Declarations

### Ethics approval and consent to participate

We obtained ethical approval from the **Busitema University Faculty of Health Sciences Research Ethics Committee (BUFHS-REC), Busitema University, Uganda** (REC approval number: **BUFHS-2023-117**) before commencement of the study. The Committee issued a letter of approval and an introduction letter authorising the conduct of the research. Administrative clearance was obtained from the Office of the District Health Officer of Mayuge District and the authorities of the selected health facilities.

The study involved a retrospective review of routinely collected maternal, newborn and child health records. Therefore, the requirement for informed consent was waived by the BUFHS-REC. All data were handled confidentially and securely, and personal identifiers were not included in the analysis dataset.

This study was performed in accordance with the principles stated in the Declaration of Helsinki (https://www.wma.net/policies-post/wma-declaration-of-helsinki-ethical-principles-for-medical-research-involving-human-subjects/).

### Consent for publication

Not applicable

### Availability of data and materials

The datasets used and/or analysed during the current study are available from the corresponding author on reasonable request.

### Author Contributions

HE conceived the idea, HE, YL drafted part of the manuscript, FO, RN BTM performed the analysis and reviewed the manuscript, EK, RM, PT, AK, DO, MWM drafted part of the manuscript and interpreted results, DK and DM supervised the project implementation, reviewed and drafted the subsequent versions of the manuscript. All authors have read and agreed to the final version of the manuscript.

## Acknowledgement

We would like to thank all staff at the different health facilities who made the data collection process possible.

